# Automatically Identifying Event Reports of Workplace Violence and Communication Failures using Large Language Models

**DOI:** 10.1101/2024.09.18.24313893

**Authors:** Mike Becker, Sy Hwang, Emily Schriver, Caryn Douma, Caoimhe Duffy, Joshua Atkins, Caitlyn McShane, Jason Lubken, Asaf Hanish, John D. McGreevey, Susan Harkness Regli, Danielle L. Mowery

## Abstract

Safety event reporting forms a cornerstone of identifying and mitigating risks to patient and staff safety. However, variabilities in reporting and limited resources to analyze and classify event reports delay healthcare organizations’ ability to rapidly identify safety event trends and to improve workplace safety. We demonstrated how large language models can classify safety event report narratives as workplace violence and communication failures as a first step toward enabling automated labeling of safety event reports and ultimately improving workplace safety.

## Introduction

In service of Penn Medicine’s goal to become a High Reliability Organization, we aim to develop informatics solutions for analyzing trends in employee-submitted safety event reports. Event reports contain free-text narrative of the event or near-miss. Improving the ability to conduct thematic mapping and common-cause analysis of safety report data can help identify reasons and characteristics of safety incidents to target improvement efforts. Safety event reporting forms a cornerstone of identifying and mitigating risks to patient and staff safety^1^. However, due to variabilities in reports and lack of standardized processes for analyzing safety events, valuable data can be missed leading to a loss for learning and improvement^2-3^. Across safety-critical industries, natural language processing (NLP) has demonstrated significant potential in analyzing incident reports^4-6^. This technology can efficiently process vast amounts of unstructured text data from patient safety reports to yield actionable insights. NLP techniques are already effective in classifying incidents by type, medication errors, or harm severity, showing promising performance outcomes^7-10^. Despite these advances, methodological challenges persist in generalizing these results. To fully realize NLP’s potential in enhancing patient safety analyses, we must address the unique challenges of applying NLP to healthcare narratives.

Previously, we assessed the feasibility of identifying event reports about violence using NLP techniques such as Gradient Boosting, Logistic Regression, and Random Forest, with the highest performance achieved by Extreme Gradient Boosting (AUROC: 0.806; APS: 0.721) on the validation set, and comparable performance observed on the development set^11^. While this performance was promising, we aim to improve upon this initial pilot by incorporating large language models (LLMs), a state-of-the-art advancement in natural language processing (NLP), and assessing their ability to interpret and categorize narratives from the same data set. The primary objectives of this study were to evaluate the performance of LLMs in automatically classifying the topic of safety event reports according to workplace violence and communication failures. This paper describes an annotation and automation study conducted to create a coded dataset for use in applying an n-shot learning approach with LLMs.

## Methods

This retrospective, observational study was approved by the University of Pennsylvania Institutional Review Board.

### Safety Net Classifications and Codebook Development

We selected safety event reports (n=2,100 narratives) from the RLDatix safety event reporting system from July 1, 2022 through December 21, 2022. Prior to analysis, each safety event report and its associated narrative was de-identified using PHIlter^12^. When reporting a safety event, reporters are limited to only 1 category label by the safety event reporting system; however, often multiple labels may apply to a narrative. In this synthetic example, “*Patient arrived for a visit that was cancelled, and became agitated when I told them they couldn’t be seen today. They yelled at me, slammed their hand on the desk, and pushed me back when I tried to talk to them. Security was called. After the patient left, the office manager blamed me for not telling the patient that the appointment was cancelled*.”, the labels should be physical violence, verbal abuse, and communication failure. We identified all event reports with the following initial labels by an original reporter from the safety net reporting system as **violence** (n=861 narratives): Aggression toward an inanimate object, Violence/injury to visitor, Restraints-injury in restraints, Intimidation/verbal abuse, Harassment, Sexual harassment, Physical abuse, Healthcare worker violence: discrimination, Verbal abuse, Healthcare worker injury, Laceration, Healthcare worker violence, and Inconsiderate/rude/hostile/inappropriate. We also identified and preserved all event reports with initially labeled by an original reporter from the safety net reporting system as **communication** (n=245 narratives). A codebook was developed to define general categories of workplace violence and communication failures based on reporting utility and prior works^13^.

### Communication categories included

- **Accurate communication:** How accurate is their communication with each other? If information is not accurate it may lead to errors and delays, and may influence future knowledge seeking. Did inaccurate information contribute to this event?
- **Frequent communication:** How frequently do people in each of these groups communicate? The frequency of communication between participants.
- **Problem solving communication:** Where there is a problem, do the people in these groups blame others or try to solve the problem? Do they blame each other when errors occur causing information to go underground rather than being shared? Blame is important to qualify for **Problem solving communication** code.
- **Timely communication:** How timely is their communication with each other? Are there delays or is it punctual? Could this event have been impacted by earlier communication? Note: does not apply if the problem arose from just a technical issue; for example, overhead rapid response was called but no phone call was received.

### **Violence** subcategories included

- **Physical violence:** Did an act or threat occur at the workplace that can include any of the following: nonverbal, or physical aggression; threatening, intimidating, harassing, or humiliating actions; bullying; sabotage; sexual harassment; physical assaults; or other behaviors of concern involving staff, licensed practitioners, patients, or visitors? Can include violence toward an inanimate object (for example, breaking a window).
- **Verbal abuse:** Did an act or threat occur at the workplace that can include any of the following: verbal, written aggression; threatening, intimidating, harassing, or humiliating words; bullying; sabotage; sexual harassment; physical assaults; or other behaviors of concern involving staff, licensed practitioners, patients, or visitors?
- **Verbal threat of future violence:** Was a threat of violence or aggression expressed including a direct or implied physical threat such as fighting or coming back to hurt someone or a direct or implied threat of non-physical action such as reporting or suing? Threat only applies to a verbal action; if someone picks up an object and starts to throw it but stops, that is still coded as **Physical violence**.

### Non-codeable

Does the report does not fit in any of the above categories? Code will be used to verify that the report has been reviewed.

### Annotation Study

Each safety event narrative was independently annotated by three clinical annotators (CD, JA, SR) with one or more of the aforementioned labels. A reference standard of labels was determined through consensus review; the codebook was updated following each batch of review (n=6 rounds). We assessed the agreement between three annotators for coding safety event reports (n=314 narratives) with labels according to **violence** and **communication failure** categories and *subcategories*. We computed inter-annotator agreement (IAA) as positive agreement (ppos) across annotator pairs (e.g., A1/A2, A2/A3, A1/A3), full agreement (all A# agree), and any two pairs (e.g., A1/A2 or A2/A3 or A1/A3). We also report the number of true positives (tp), true negatives (tn), and false positives + false negatives (errors) for the final batch reviewed (round 6).

### Automation Study

To automatically classify safety event report narratives, we leveraged OpenAI’s GPT-4o model, which has shown promising results on tasks involving medical narratives. To ensure reproducibility, we kept parameters static with temperature at 0.1 and top_p at 1. We then instructed the model to classify one or more categories of **violence** or **communication failures** for each narrative. We utilized a three-part prompting strategy consisting of a task prompt, instruction prompts for each subcategory, and a system prompt to guide the model’s classification tasks. We iteratively refined the task and instruction prompts for classifying narratives using subset of the dataset for development.

The task prompt we gave GPT-4o provides context and a role to apply each of the subcategory labels:

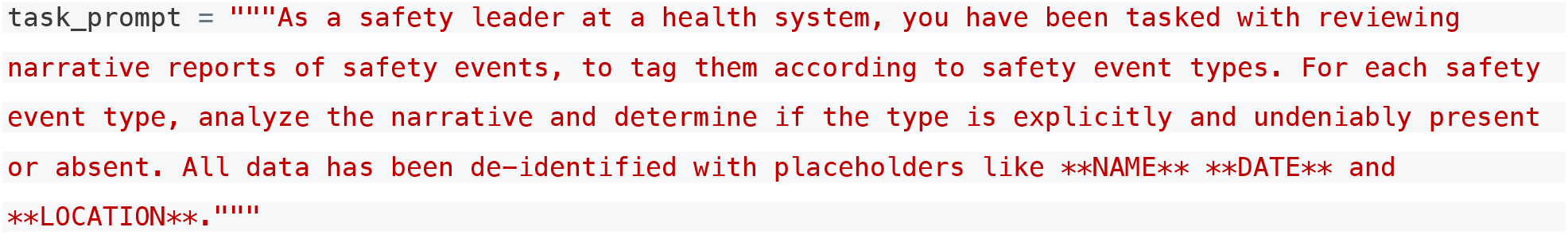

Next, we provided the following instruction prompts including examples for each subcategory:

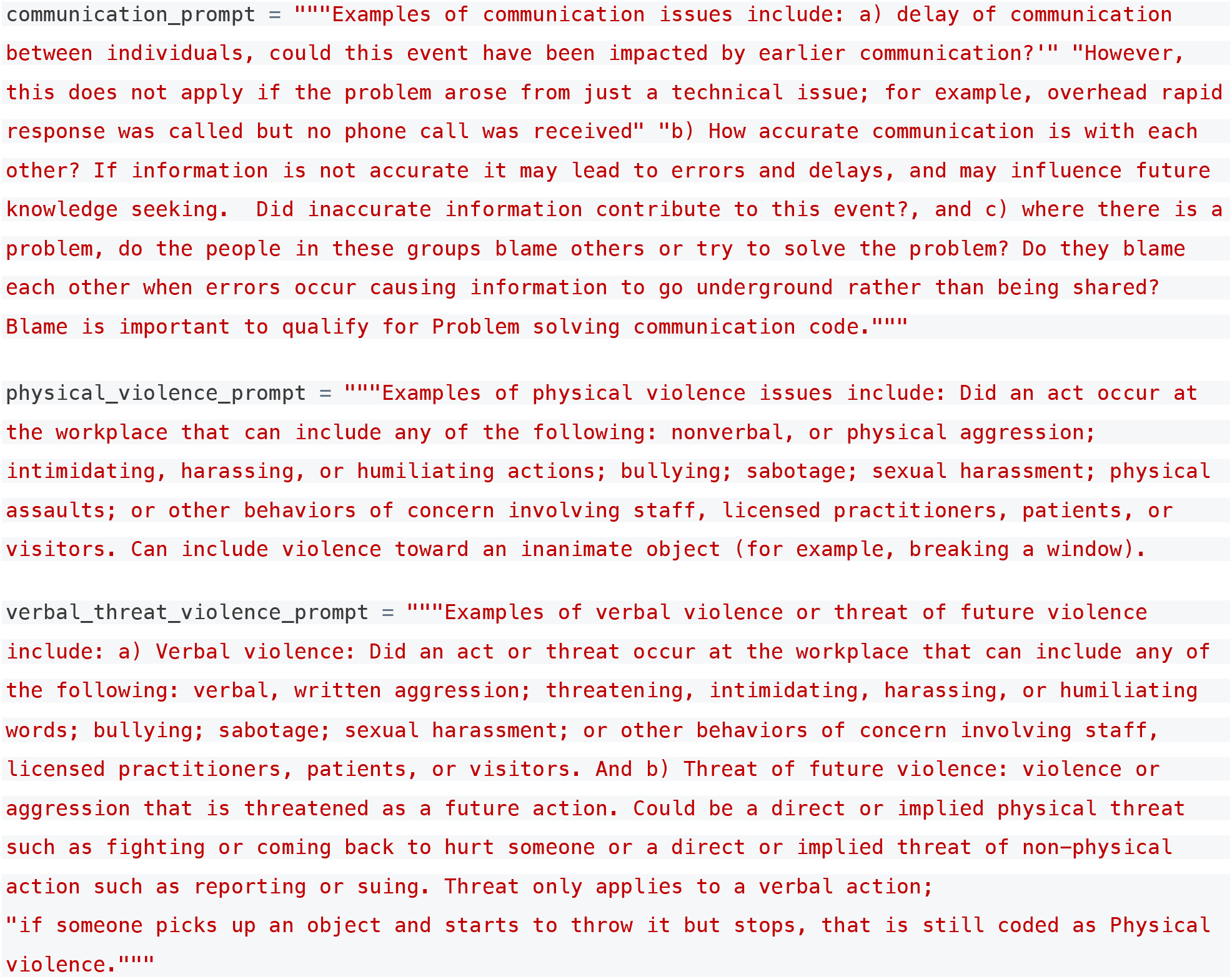

Lastly, this system prompt was designed to ensure that the model’s output is structured in a consistent and machine-readable format, facilitating seamless integration with our downstream processing pipelines.

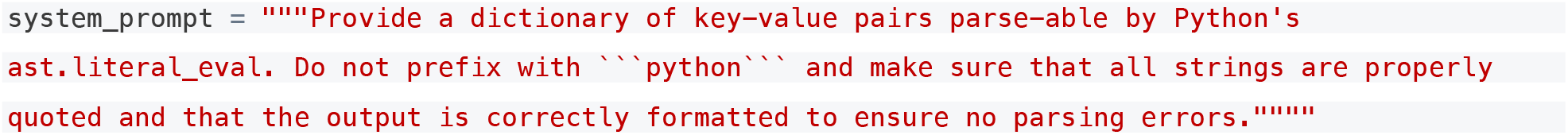

We report performance for the dataset including precision, recall, and F1-score (the harmonic mean of precision and recall) for **violence** and **communication** *subcategories*^13^.

### Relationship between Classifier Performance with Annotator Agreement and Certainty Levels

We then conducted subanalyses to understand the relationship between classifier performance and both annotator agreement (minimal threshold levels) and certainty levels (exact agreement levels). Specifically, for each narrative, we encoded the number of annotators that agreed with the resulting reference standard label as a proxy to certainty and difficulty in assigning a label, i.e., narratives in which all three annotators agreed with the resulting label might exhibit absolute agreement with high certainty; in contrast, narratives in which all three annotators did not agree with the resulting label might exhibit no agreement and no certainty.

Therefore, we characterize this in two ways compared to the resulting reference standard label:

- Minimal Threshold Levels (at least N annotators agreed with the reference standard): 3 annotators (*absolute agreement*), 2+ annotators (*majority agreement*), 1+ annotators (*any agreement)*, 0 annotators *(no agreement)*
- Exact Agreement Levels (exactly N annotators agreed with the reference standard): 3 annotators only (*high certainty*), 2 annotators only (*moderate certainty*), 1 annotator only (*low certainty*), 0 annotators *(no certainty)*

For each category and subclass, we report classification performance for subsets of the dataset demonstrating different definitions of annotator agreement and certainty levels.

### Random vs Ordered N-Shot Learning Examples

In order to optimize performance, we applied an n-shot learning (n=0, 2, 6, 10; equal positive and negative examples) approach. However, we wanted to understand the impact of providing well-detailed versus more ambiguous training examples for instruction prompting. Training examples were ranked by clarity (well-detailed examples were characterized as having complete details of the story and containing words and phrases associated with the category with high confidence). To evaluate example ordering on instruction prompting, we used the same set of randomly selected n-shot examples for both comparison arms. Then, we created two arms of comparison, *random* versus *ordered* n-shot learning examples. In *random*, we randomly selected the order of examples provided within the prompts; in *ordered*, we presented the same examples in a fixed sequence prioritizing based on clarity. We hypothesized that the order in which examples are presented may influence initial performance, with earlier examples that are more direct and less ambiguous potentially having a greater impact compared to random selection of labeled training examples.

## Results

We conducted an annotation and automation study of safety event report narratives.

### Annotation Study

We assessed the agreement between three annotators for coding safety event reports (n=314 narratives) with **violence** and **communication** categories and *subcategories*. We computed inter-annotator agreement (IAA or positive agreement) across annotator pairs, full agreement, and any two pairs; we report the last batch (round 6) for agreement (**Figures 1** and **2**). In **Figure 1**, where a majority of annotators agreed (any 2), *not codeable* is the most frequent category (n=45 tp) followed by *verbal abuse/verbal threat of future violence* (n=26 tp) and *physical violence* (n=18 tp). In terms of overall IAA across rounds (any 2) for **violence**, we observed IAA ranging from 0.87 (round 3) to 0.97 (round 1). More specifically, for *physical violence*, we observed IAA ranging from 0.00 (round 4) to 0.92 (round 6). For *verbal abuse/verbal threat of future violence*, we observed IAA ranging from 0.87 (round 6) to 1.00 (round 1). After round 5, we observed IAA just at or greater than 0.50 for all (sub)categories.

**Figure 1.**
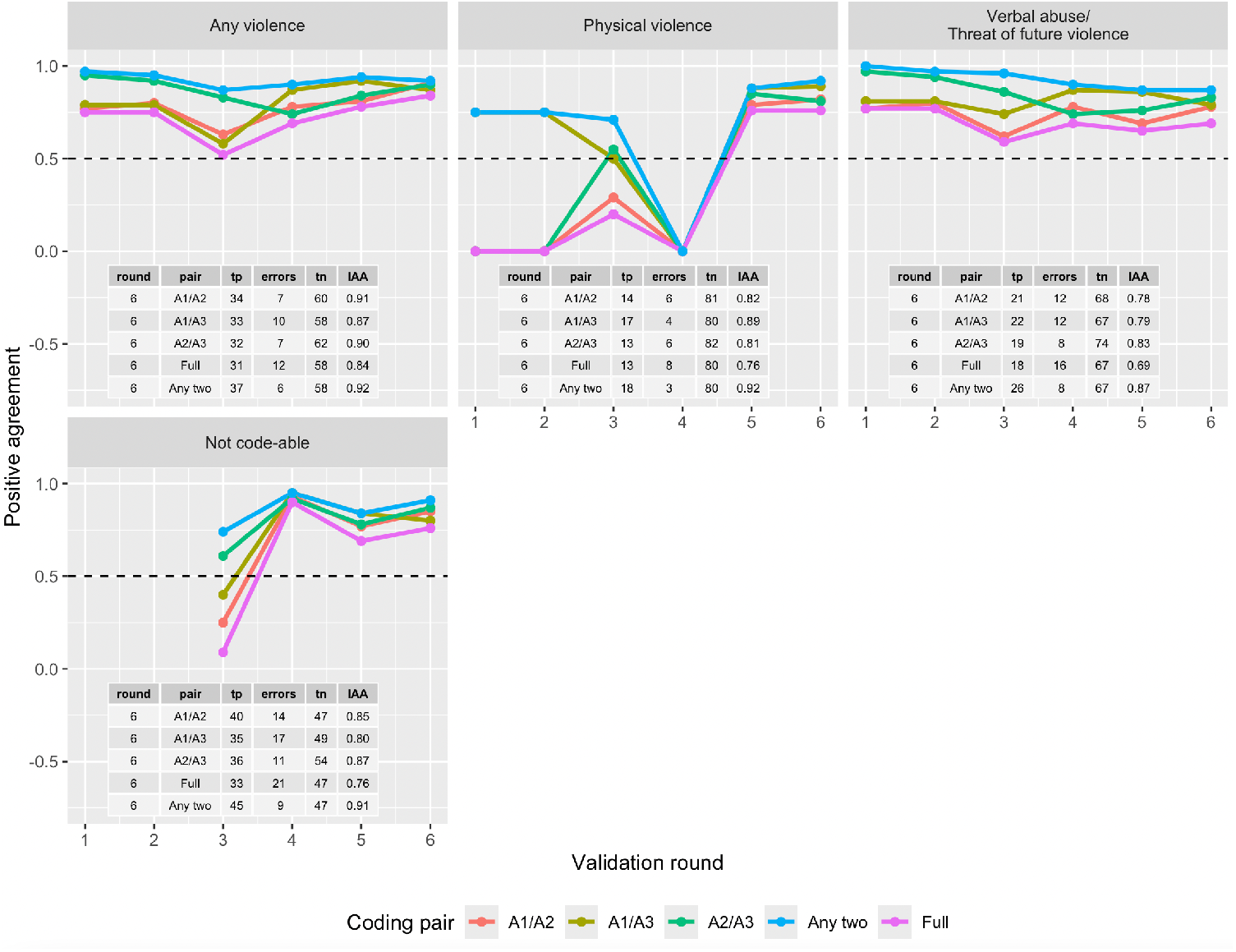
IAA across annotator pairs, full, and any two annotators for violence categories and subcategories. IAA reported across rounds (batches). Rounds enumerated along the x-axis; IAA (positive agreement) listed along the y-axis from 0 to 1.

**Figure 2.**
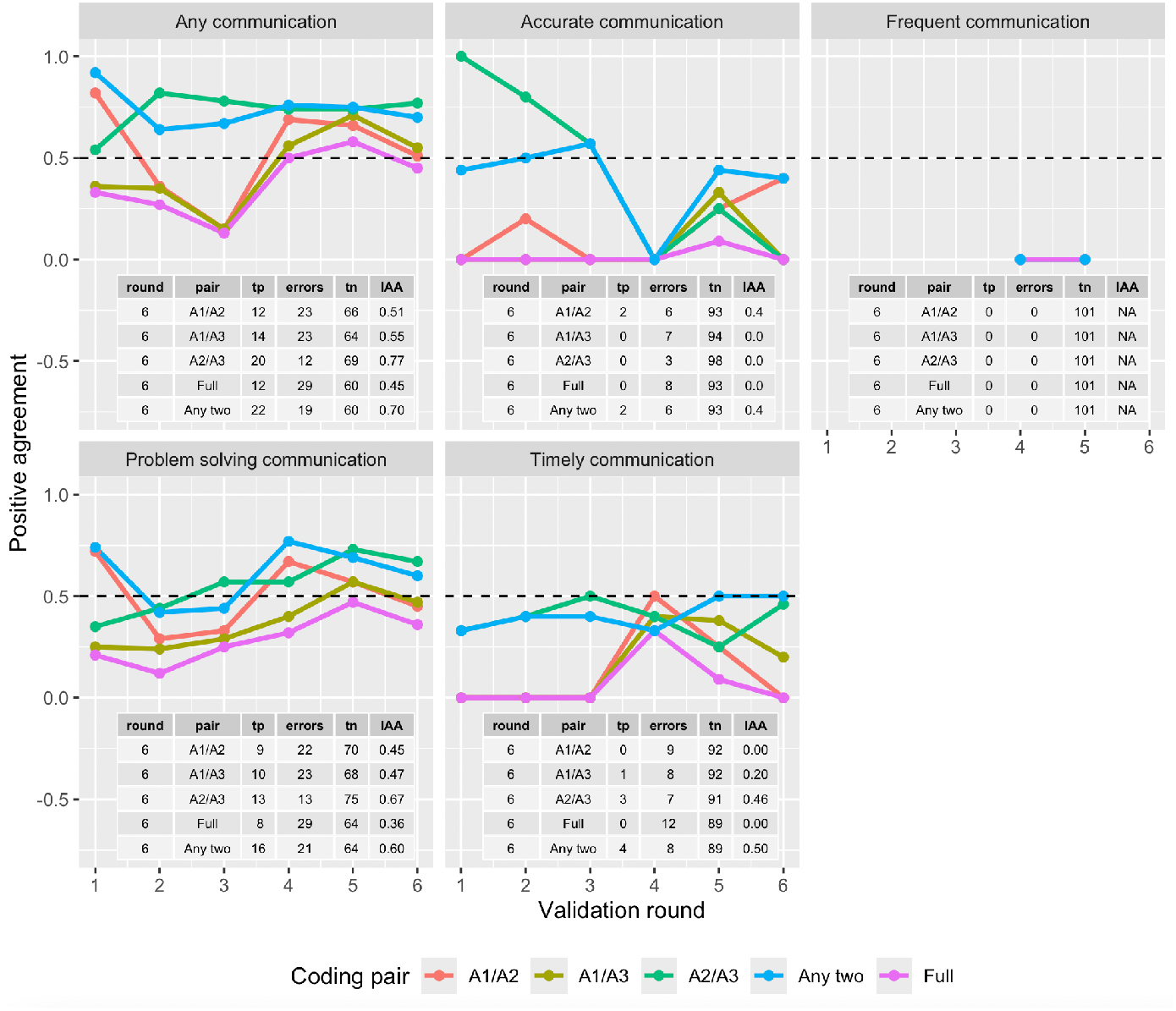
IAA across annotator pairs, full, and any two annotators for communication failures categories and subcategories. IAA reported across rounds (batches). Rounds enumerated along the x-axis; IAA (positive agreement) listed along the y-axis from 0 to 1.

In **Figure 2**, where a majority of annotators agreed (any 2) in round 6, *not codeable* is the most frequent category followed by *problem solving communication* (n=16 tp), *timely communication* (n=4 tp) and *accurate communication* (n=2 tp). In terms of overall IAA across rounds (any 2) for *any* **communication**, we observed IAA ranging from 0.64 (round 2) to 0.92 (round 1). After round 4, we observed IAA at or greater than 0.50 for *problem solving communication* and *timely communication* only.

### Automation Study

Due to low IAA across some subcategories, we collapsed all subcategories of **communication** into one category. Similarly, we collapsed *verbal abuse* and *threat of future violence* into one subcategory and maintained *physical violence* as a subcategory. This resulted in three categories for classification. In **Figure 4**, in terms of overall performance according to annotator agreement levels of *absolute, majority*, and *any agreement*, we observe F1-scores ranging from 0.67 to 0.94. We then conducted subanalyses to understand the relationship between classifier performance and both minimal agreement and annotator certainty. For **communication**, the best classifier performed at F1-score of 0.94 for narratives with absolute agreement using random ordering with 2-shot learning. Consistently, 2-shot learning performed best for both random and ordered examples. For *physical violence*, the best classifier achieved an F1-score of 0.80 across all agreement levels using random ordering with 6-shot and 10-shot learning. For *verbal abuse/threat of future violence*, the best classifier performed at F1-score of 0.94 for narratives with absolute agreement using random ordering with 2-shot learning. More detailed performance measures can be found in **Figure 6** in the **Appendix**.

In **Figure 5**, in terms of overall performance for certainty levels of *high, moderate*, and *low agreement*, we observe F1-scores ranging from 0.0 to 1.0. For **communication**, the best classifier performed at F1-score of 0.94 for narratives with high agreement using random ordering with 2-shot learning. For *physical violence*, the best classifier achieved an F1-score of 0.86 across all agreement levels using random ordering with at least 6-shot learning. For *verbal threat of violence/threat of future violence*, the best classifier performed at F1-score of 1.0 for narratives with absolute agreement using random and ordered training examples across 0 through 10 shot learning. More detailed performance measures can be found in **Figure 7** in the **Appendix**.

## Discussion Annotation Study

In general, annotators were able to achieve higher IAA for **violence** categories than **communication** categories. We have several explanations for this observation. First, violence-labeled narratives tend to have clear words and phrases, e.g., “attack”, “verbally abused”, “aggressive/threatening behavior”, and “yelling” within the text. **Violence**-related narratives tend to contain most pertinent information necessary to assert a **violence** category or subcategory; in contrast, **communication** failures tend to be less well described and often pertinent details are entered following the initial reports resulting in a limited information necessary to assign more descriptive **communication** categories. However, in spite of these limitations, across all categories, most safety event report narratives received the same label by the majority of annotators (2+ annotators). Distinctions among subcategories of communication and between *verbal abuse* and *threat of future violence* were often not clear; we therefore collapsed these subcategories into two larger categories: *communication* and *verbal abuse/threat of future violence* for automation.

In **Figure 3**, annotators labeled 81 *communication failures*, 42 *physical violence*, 98 *verbal threat of violence/threat of future violence*, and 135 *not codeable* narratives. Across all categories, most safety event report narratives received the same label by the majority of annotators (2+ annotators). In a rare case, we observed no agreement among annotators for narratives deemed as *not codeable* (n=8 narratives).

**Figure 3.**
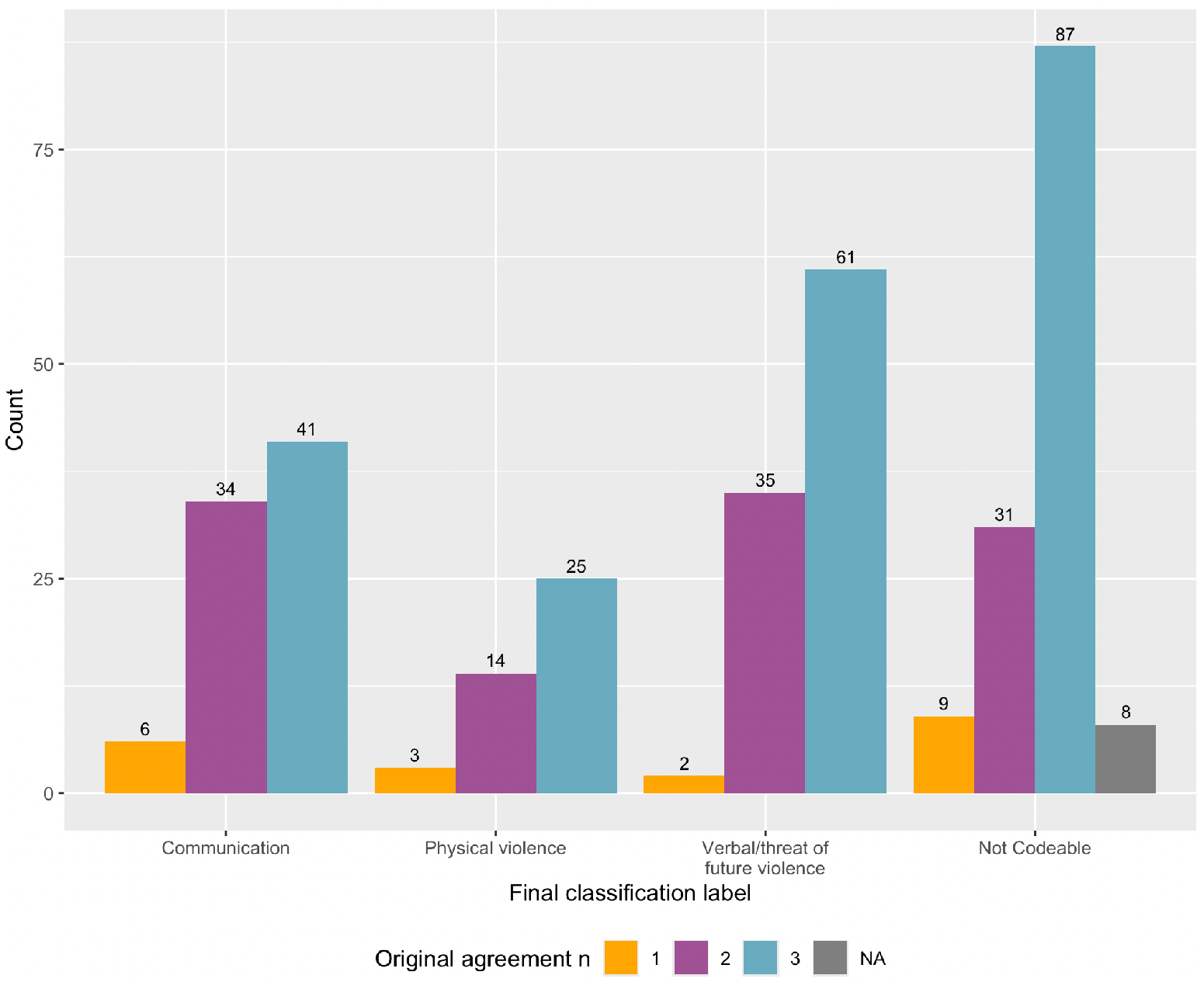
IAA across annotator pairs, full, and any two annotators for communication failures and violence categories and subcategories.

### Automation Study

In **Figure 4**, applying n-shot learning on the full dataset, we observe the highest F1-score performance using random ordering with variable n’s of learning across categories: *communication* with 2 shot at 0.90, *physical violence* with 6 and 10 shot at 0.80, and *verbal abuse/threat of future violence* with 2-shot at 0.93. In terms of minimal threshold levels as absolute (3 annotators) majority (2+ annotators), and any (1+ annotators) agreement, the highest achieved performance was observed with 3 annotators across all categories with the exception of ordered *communication* and random *physical violence* which had tied performance with 2+/3 and all datasets, respectively. However, the performance was often a marginal increase suggesting that LLMs can handle narratives which can pose to be more challenging for annotators and maintain similar performance to those with absolute agreement.

**Figure 4.**
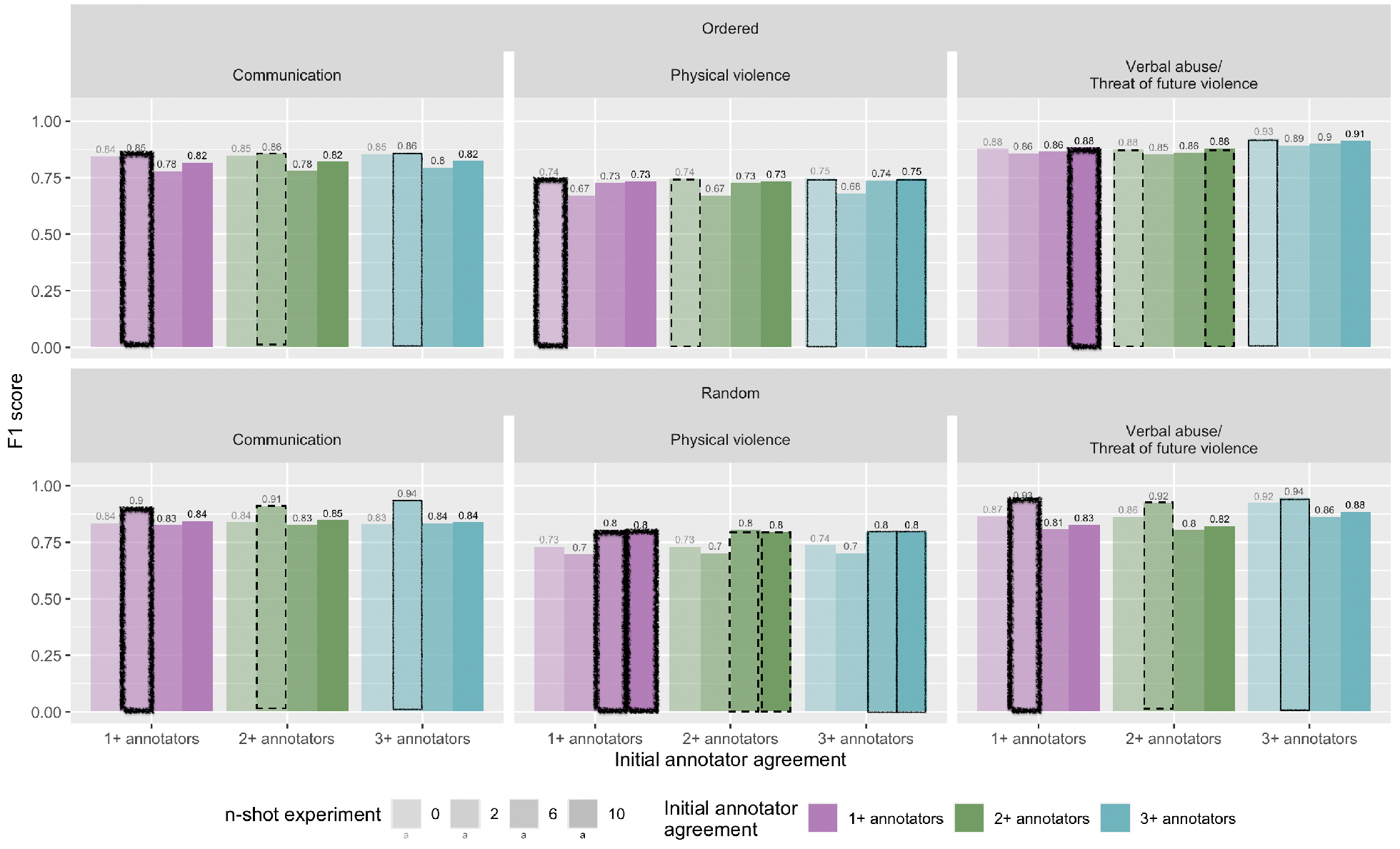
Performance of the best n-shot learning according to F1-score positive class per category by annotator agreement levels defined as *absolute, majority*, and *any agreement* with the reference standard. Each set of 4 continuous bars of same color represents n-shot learning experiments within a category and minimal threshold agreement dataset. Highlighted bars indicate the highest performing classifier within that experiment.

In **Figure 5**, applying n-shot learning and varying certainly levels as high (3 annotators), moderate (2 annotators only), and any (1 annotator only) agreement, we observe the highest performance using random ordering with variable n’s of learning across categories: *communication* with 2-shot at 0.88, *physical violence* with 6-shot at 0.86, and *verbal abuse/threat of future violence* tied random and ordered with all-shots at 1.0. In terms of certainty levels, it’s surprising that random performed well. It could be that some more clear examples were still randomized and introduced early in ordering. Another hypothesis is that what might be clear to a human might not provide the same informativeness to the machine. It’s worth reiterating that generative AI in healthcare is an emerging application area and the natural stochasticity of LLMs can lead to inconsistent and non-intuitive performance. Also, in the case of *verbal abuse/threat of future violence* highest performance was observed for those cases with low agreement (1 annotator only). While that observation is surprising, given we only observed two instances of low agreement we do not believe it’s wise to draw any conclusions until we have implemented this on a larger dataset.

**Figure 5.**
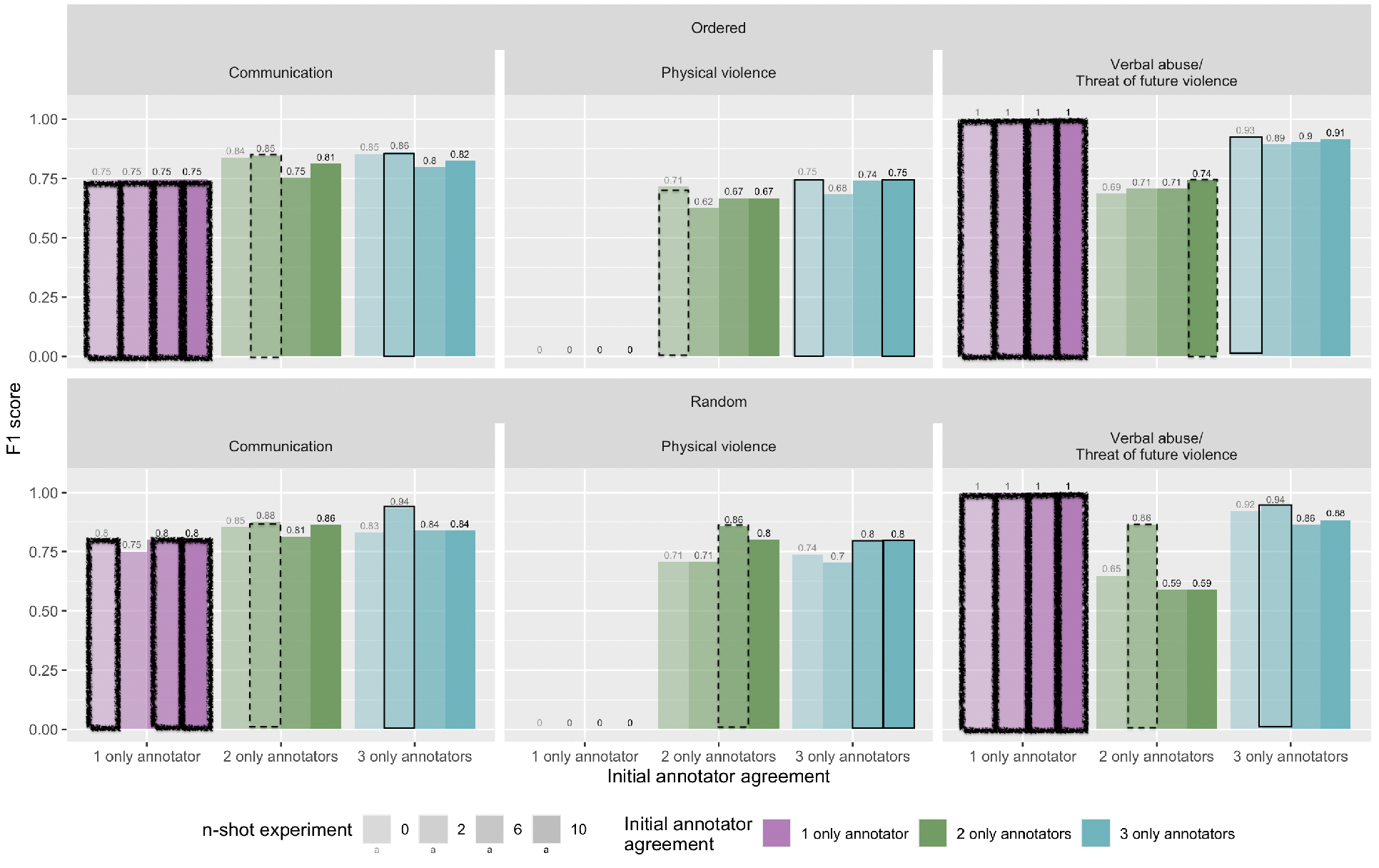
Performance of the best n-shot learning according to F1-score positive class per category by certainty levels defined as *high, moderate*, and *low agreement* with the resulting reference standard. Each set of 4 continuous bars of same color represents n-shot learning experiments within a category and exact agreement levels dataset. Highlighted bars indicate the highest performing classifier within that experiment.

### Limitations and Future Directions

This study has several limitations. First, we only reviewed and automated safety event report narratives from one institution and across a 6-month period. Narratives across multiple institutions and more writers may generate different performance. However, given the size of GPT-4o we suspect performance will be fairly consistent with new narratives. We did not algorithmically determine which example labeled safety event report narratives would iteratively improve performance. In the future, we will implement and evaluate advanced retrieval-augmented generation (RAG) techniques to continually enhance the performance of LLMs in clinical text analysis and sample the most informative examples defined by the LLM (rather than humans) over time. Additionally, we will experiment with fine-tuning strategies for LLMs to further optimize the classification of narratives across categories. Lastly, we only automated two types of safety event reporting categories. We will future expand our efforts to include new categories.

## Conclusion

We demonstrated how large language models can automatically classify safety event report narratives as workplace violence and communication failures with high reliability as a first step towards improving automatic data labelling of safety event reports.

## Data Availability

Although, all data were de-identified prior to analysis due to the sensitivity of the subject matter and patient re-identification risk, these data are not available.

## Acknowledgements

We thank the Center for Applied Healthcare Informatics and Penn Medicine leadership for their support of this translational (research to operations) project.

## Appendix

**Figure 6.**
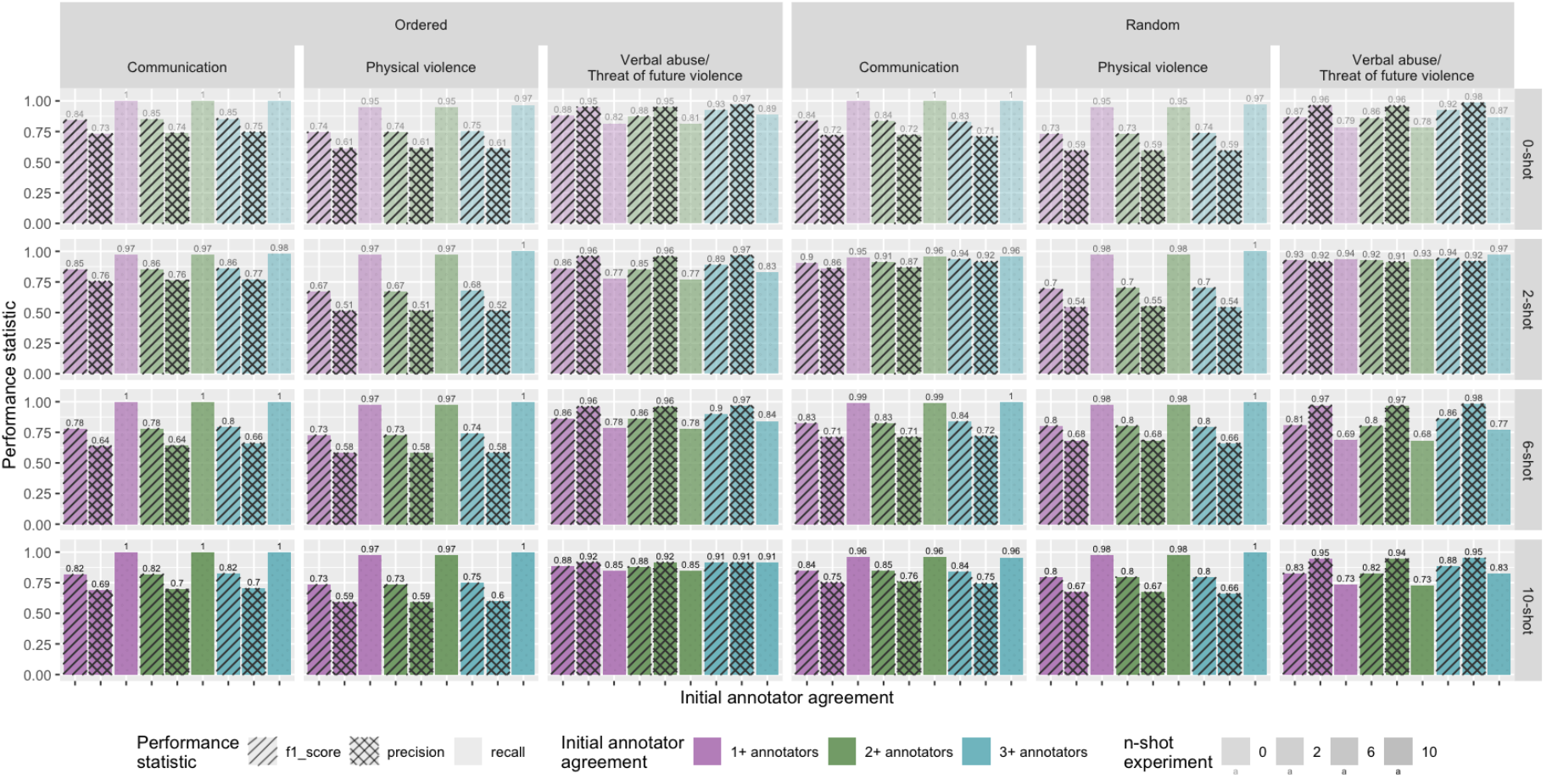
Performance of the best n-shot learning according to F1-score, precision and recall for the positive class per category by annotator agreement levels defined as *absolute, majority*, and *any agreement* with the reference standard.

**Figure 7.**
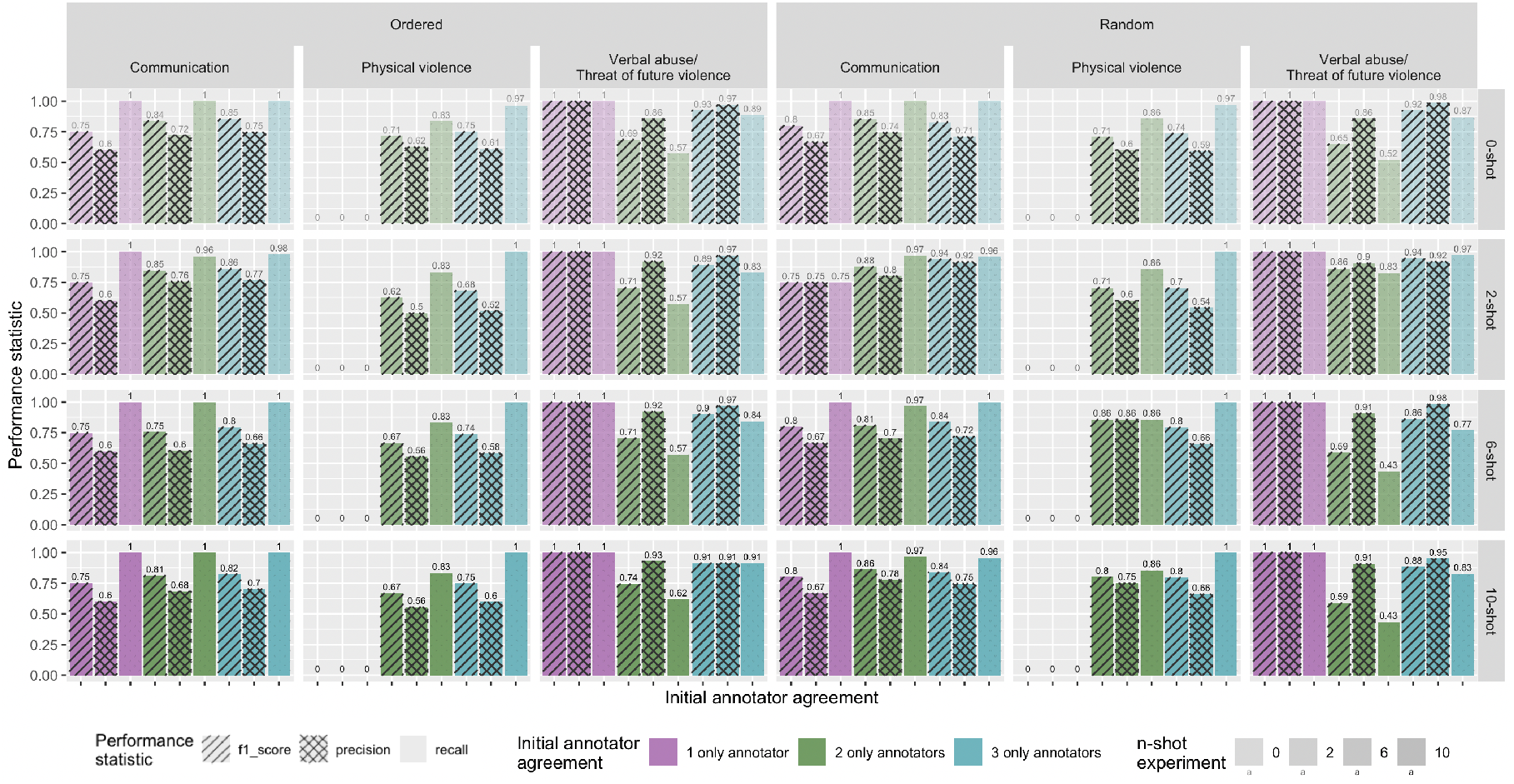
Performance of the best n-shot learning according to F1-score, precision, and recall positive class per category by certainty levels defined as *high, moderate*, and *low agreement* with the reference standard.

## Notes

### Competing Interest Statement

The authors have declared no competing interest.

